# Optimization and clinical validation of new and improved TaqMan Real-Time PCR assays for the detection of pathogenic *Leptospira*

**DOI:** 10.64898/2026.08.13.26359137

**Authors:** Camila Hamond, Alice Zhao, Luiza Aymée, Walter Lilenbaum, Ilana T. Balassiano, Elsio A. Wunder

## Abstract

Leptospirosis is an infectious neglected zoonotic disease caused by pathogenic bacteria of the genus *Leptospira*. The genus comprises 43 pathogenic species, divided into two clades (P1 and P2), with the potential to cause disease on animals and humans. Despite the major impact of this disease on animal and human health, few quantitative real-time polymerase chain reaction (qPCR) assays have been validated to specifically detect all pathogenic *Leptospira* species, thwarting diagnosis and epidemiological studies. The gene encoding LipL32, the major leptospiral outer membrane protein, discriminates pathogenic P1 species from P2 and saprophytic. However, with the recent discovery of new species, the current *lipL32*-based qPCR assay cannot detect all classified P1 species. Furthermore, there are no currently validated molecular methods able to differentiate the presence of P1 and P2 species on clinical samples. Previous analyses have shown that the *23S ribosomal RNA* gene displays considerable conservation in P1 and P2 species but sequence divergence in saprophytic species, a promising target for PCR-based detection and discrimination of those two clades. This study optimized and validated an improved *lipL32*- and *23S*-based TaqMan qPCR assay using human and animal clinical samples. These newly optimized and developed assays resulted in a lower limit of detection and increased diagnostic sensitivity, resulting in the detection of all pathogenic species of the genus *Leptospira* currently described. These assays will improve the detection of leptospires from clinical and environmental samples, providing a valuable epidemiological and clinical tool to support One Health research on this important emerging disease.

**IMPORTANCE:** Leptospirosis is neglected disease. Detection of leptospiral antibodies through a microscopic agglutination test (MAT) remains the gold standard for leptospirosis diagnosis. However, MAT, culture and other serological methods, such as ELISA and rapid test, have inconsistent sensitivities and limited benefit for early diagnosis of leptospirosis. Early diagnosis of leptospirosis is important for ensuring better clinical management and achieving better outcomes. qPCR is more sensitive, specific and able to detect the presence of pathogenic *Leptospira* DNA in clinical samples. With the recent discovery of new leptospiral species, the sensitivity of the current *lipL32*-based qPCR assay has been affected unable to detect all currently identified P1 and P2 species. To address this issue, this study optimized and validated an TaqMan qPCR assay using animal and human clinical samples.

## INTRODUCTION

Leptospirosis is the most widespread zoonosis globally, caused by pathogenic spirochetes of the genus *Leptospira*. The neglected status of leptospirosis as a serious public health issue has contributed to its high morbidity and mortality, especially in resource-poor countries in tropical regions (1). As an environmentally transmitted disease, spillover infections to humans and animals occur mostly through exposure to soil or water contaminated with the urine of reservoir animals infected with pathogenic *Leptospira* (2). In humans, infection caused by pathogenic leptospires manifests a wide spectrum of clinical features, ranging from asymptomatic or mild illness to a severe life-threatening disease, with an estimated 1 million cases and 60,000 deaths annually (1, 3). Leptospirosis can also cause a life-threatening disease in domestic animals and significant economic impact on livestock (4).

Currently, the genus comprises 74 species divided into four subclades that correlate to their ability to cause disease in mammal hosts. Two subclades include 43 species considered pathogenic, 21 P1 and 22 P2, that may cause infections in animals and/or humans, as well as environmental species with uncharacterized virulence. The other two subclades include 31 species considered saprophytic, 26 S1 and 5 S2, that thrive in soil and/or water unable to cause infections in humans or animals (5–7).

Leptospires penetrate abraded skin and mucous membranes and have potential to establish systemic infection by crossing tissue barriers, resulting in hematogenous dissemination (8). During the septicemic phase, leptospires are found in the bloodstream during the first few days after exposure, followed by an immune phase which is characterized by the development of antibodies and subsequent clearance of leptospires from the bloodstream (2). Culture isolation of causative organisms from biological fluids (blood, cerebrospinal fluid, tissue, or urine) can take several weeks, and antibodies can be detectable in the blood by serological methods approximately one week after the onset of symptoms (2). Alternatively, several molecular assays have been described for leptospiral detection in clinical samples during the initial phases of the infection (9–16).

The *lipL32* gene, which encodes a leptospiral outer membrane lipoprotein, has been described as a putative virulence factor absent in saprophytic species (17, 18). This specificity for pathogenic species distinguished the *lipL32* gene as a suitable and commonly used target, including a clinically validated and widely used qPCR assay designed to identify *Leptospira* species of P1 clade (9, 13, 19). *Leptospira* species have two copies of the *23S ribosomal RNA* gene (rll1 and rrl2) (20), which displays considerable conservation in P1 and P2 species but sequence divergence in saprophytic species. The *23S rRNA* gene has been described as a promising target for PCR-based detection and differentiation between P1 and P2 species (10).

With the recent discovery of new leptospiral species (5–7), the sensitivity of the current *lipL32*-based qPCR assay has been affected—unable to detect all currently identified P1 species. Furthermore, there is a lack of clinically validated qPCR assays capable of detecting *Leptospira* species from the P2 clade. The development and validation of robust detection tools capable of identifying all pathogenic *Leptospira* species in various contexts, as well as the improvement of previously designed qPCR assays to account for more recently discovered species, is a critical need in the field. To address this issue, this study optimized and validated an improved *lipL32*- and a novel *23S*-based TaqMan qPCR assay using animal and human clinical samples.

## MATERIALS AND METHODS

### *Leptospira* strains and culture conditions

*Leptospira* strains were cultivated in liquid Ellinghausen-McCullough-Johnson-Harris (EMJH) medium at 30 °C. Leptospires were inspected weekly under dark-field microscopy. Intact motile leptospires were enumerated by dark-field microscopy and Petroff-Hausser chamber as previously described (21). *L. interrogans* serovar Copenhageni strain Fiocruz L1-130, *L. borgpetersenii* serovar Hardjo strain JB197, and *L. licerasiae* serovar Varillal strain VAR 010 were used as representatives of P1 and P2 species for spiking experiments and/or DNA extraction to determine the lower limit of detection (LLOD).

### DNA extraction

*Leptospira* cultures (5 mL) were centrifuged at 12,000 × g for 10 min at 4 °C. After two washes with phosphate-buffered saline (PBS), the supernatant was removed and the pellet was resuspended in 200 μL of PBS and extracted using the Maxwell Tissue DNA Kit (Promega), following the manufacturer’s instructions, modified by using 100 μL as elution volume (22). Each DNA sample was diluted to achieve 10^6^ copies of equivalent genomic DNA (GEq) per 5 μL of extract based on the size of each genome. Genome sizes were retrieved from the National Center for Biotechnology Information (NCBI) Genome database.

### Primers and probes design

Genome sequences from all 74 species of *Leptospira* were obtained from the NCBI Genome database (Table S1) and uploaded into the Galaxy software (23). Sequences of *23S and lipL32* were downloaded from the Microbial Genome Annotation and Analysis Platform (24) and uploaded into the Galaxy software. The “NCBI BLAST + blastn” function of the Galaxy software was used to locate the sequence of both genes for each respective species. The sequences for each strain were aligned using the BioEdit software (25). Following *23S* primer and probe design, avoiding primer-dimer or probe-dimer formation, all primer and probe sequences were analyzed for potential interactions and/or secondary structures using NET Primer analysis software by PREMIER Biosoft (26). *In silico* evaluation was performed to ensure reaction efficiency, and all primer and probe sequences were evaluated using BLASTn search of the NCBI database to confirm specificity to *Leptospira* species.

Optimization of the *lipL32* assay: The primers and probes targeting the *lipL32* gene aimed to identify samples containing all and only the current 21 P1 species. The *lipL32* gene is absent in S1 and S2 species but is present in all P1 and P2 species. We performed analysis using Primer ExpressTM Software v3.0.1 to optimize a previous primer set (27) adapted from the original 2009 qPCR assay (9).

Development of the *23S* assay: The goal was to develop and validate a *23S rRNA*-based multiplex assay designed to simultaneously detect and distinguish between P1 and P2 clades of *Leptospira* species. The *23S rRNA* sequence (LIC 10928 from Fiocruz L1-130 and AHOO02_rRNA1 from VAR10) was selected based on previous analyses that identified this gene as a region that could successfully discriminate between pathogenic and saprophytic *Leptospira* species (10). A target sequence for the development of probes specific to P1 and P2 species was selected (position 1509-1528) with high variability between the P1 and P2 groups but high conservation within species of each group. Regions flanking this target sequence were analyzed for primer optimization, focusing on regions with conserved sequences in P1 and P2 species and enough divergence from saprophytic species to avoid amplification of the latter. We designed a single pair of primers able to amplify sequences from both P1 and P2, while the probes were designed for individual identification of P1 and P2, and conjugated with VIC/HEX and FAM fluorophore, respectively.

### Real-Time PCR assay

*lipL32* assay: The reaction mixture was composed of 12.5 μL of Fast Advanced Master Mix (Applied Biosystem), 500 nM of forward and reverse primer, 100nM of probe, 1 μL of VetMAX Xeno Internal Positive Control (IPC) - VIC Assay (Life Technologies), 5 μL of DNA, and as needed ultrapure water or 5 mg/mL of Bovine Serum Albumin (BSA) for a total volume of 25 μL (Table 1). qPCR was performed using a QuantStudio™ 3 Real-Time PCR System (Life Technologies). The assays were subjected to conditions of 2 minutes at 95 °C, and 40 total cycles of amplification (15 seconds at 95 °C and 1 minute at 60 °C). To control Real-Time PCR inhibitors, IPC was added to the master mix to confirm DNA amplification, detect false negatives, and to qualitatively detect presence of amplification inhibitory substances in a sample. If an IPC result was negative (inhibitors), samples were diluted 1:10 and Real-Time PCR repeated. If the diluted sample was still negative for IPC, a new DNA extraction was performed.

**Table 1:**
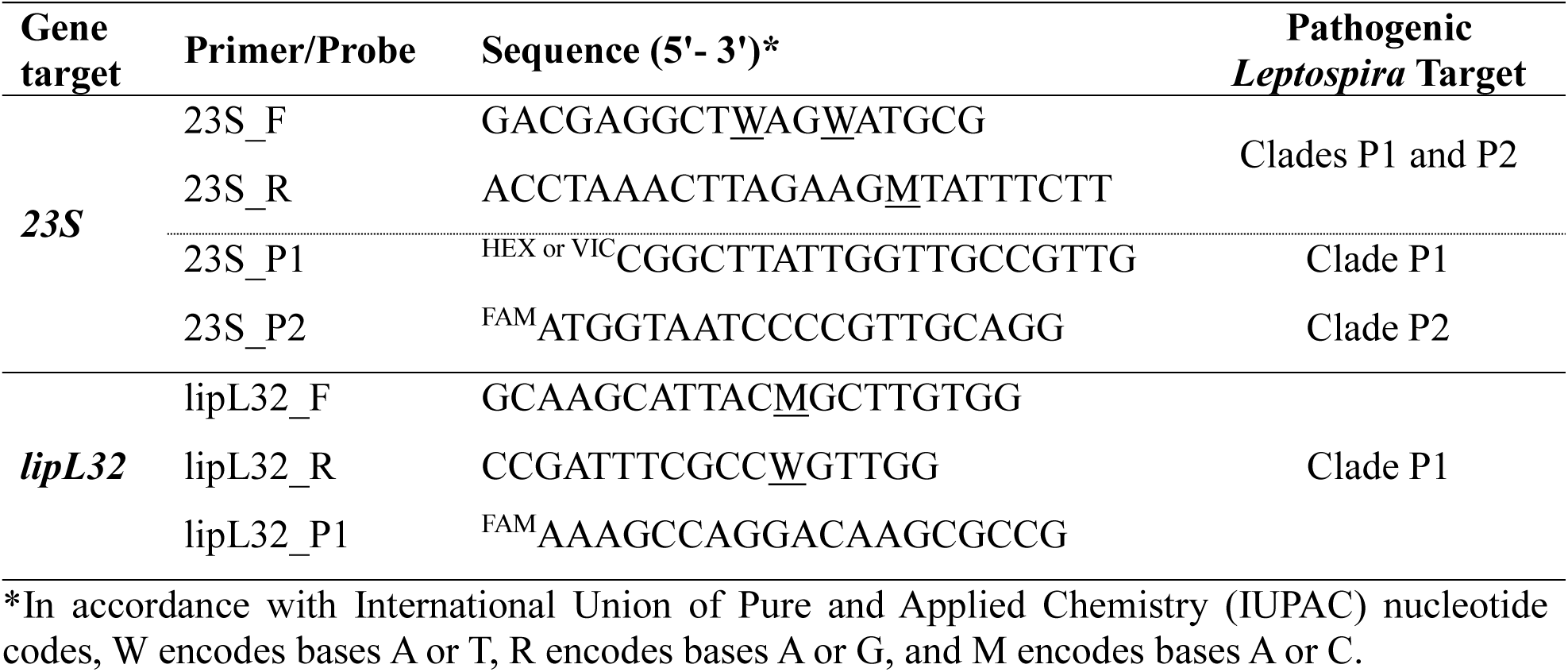
Primer and probe sequences designed and/or optimized for qPCR-based detection of pathogenic *Leptospira* species.

*23S* assay: When assessing the efficacy of the multiplex assay targeting *23S rRNA*, the sensitivity of each respective probe was analyzed individually and compared to the multiplex assay. After testing various primer and probe concentrations, the lowest concentrations that still retained for high reaction efficiency were selected. The multiplex reaction mixture was composed of 12.5 μL of Fast Advanced Master Mix (Applied Biosystem), 500 nM of forward and reverse primer, 100 nM of each respective probe, 5 μL of DNA, and as needed ultrapure water or 5mg/mL of Bovine Serum Albumin (BSA) for a total volume of 25 μL (Table 1). The assays were subjected to conditions of 2 minutes at 95 °C, and 45 total cycles of amplification (15 seconds at 95 °C and 1 minute at 54 °C). The IPC was not included in the 23S assay since it was already present in the *lipL32* assay, and the VIC dye is part of one probe of the *23S* assay. If the assays are done individually, an IPC option for *23S* assay would be the VetMAX Xeno Internal Positive Control (IPC) - LIZ (Life Technologies).

All samples were tested in duplicates and considered positive when duplicates were positive with *Ct* value < 40 and < 45 for *lipL32* and *23S*, respectively. If we observed differences in the duplicates, the sample was repeated and considered positive if both or one duplicate was still positive.

### Calculation of Analytical Sensitivity and PCR Efficiency using Lower Limit of Detection (LLOD)

The analytic sensitivity of the assays was determined with genomic DNA extracted from *L. interrogans* serovar Copenhageni strain Fiocruz L1-130 (P1) for *lipL32* and *23S* assay and *L. licerasiae* serovar Varillal strain VAR 010 (P2) for *23S* assay. A genome size of 4.6 Mb (28) for L1-130 and 4.28 Mb (29) for VAR 010 was used to determine the genomic equivalent (GEq) concentration per microliter of the purified DNA. To generate a standard curve, serial dilutions of the DNA were made starting at 1 × 10^7^ GEq to 1 × 10^0^ GEq/5 μL. Standard curve assays were performed in duplicate. The analytic sensitivity of the assay was calculated, and the lower limit of detection (LLOD) was determined to be the lowest sample concentration that can be reliably quantified by the reaction assay at 95% confidence. For *lipL32* assay, LLOD was determined with and without IPC.

The LLOD was defined as the concentration (copies of the gene/μL) that yielded 95% positive results along with the 95% confidence interval (CI) and determined using a Probit regression analysis implemented in the MedCalc Statistical Software version 19.0.7 (MedCalc Software bvba, Ostend, Belgium 2019).

### Spiking Experiments

Urine: bovine urine, negative for leptospires, was spiked with *L. borgpetersenii* serovar Hardjo strain JB197 (P1) to optimize parameters for urine storage and pre-processing as previously described (22). In brief, 1 mL of strain JB 197 at a density of 10^8^ leptospires/mL was inoculated into 9 mL of freshly collected bovine urine. A 10-fold dilution in urine was performed to obtain spiked samples containing 10^7^ - 10^0^ *Leptospira*/mL of urine (30). Dilutions were then processed for DNA extraction after storage in various conditions/time points, including: (A) same day, (B) same day after centrifugation at 900 × g for 10 min to remove potential debris, (C) after storage for 24 h in a Styrofoam container with ice packs, (D) after storage for 24 h in a Styrofoam container with ice packs followed by centrifugation to remove potential debris, (E) after storage for 48 h in a Styrofoam container with ice packs, (F) after storage for 48 h in a Styrofoam container with ice packs followed by centrifugation to remove potential debris, (G) after storage for 48h without ice, (H) after freezing at −20 °C for 24 h, and (I) after centrifugation at 900 × g to remove potential debris and frozen at −20 °C for 24 h. After each treatment, urine was centrifuged at 12,000 × g for 30 min. The supernatant was removed, and the pellets were washed once by resuspending in 1 mL phosphate buffered saline (PBS) and centrifuged at 12,000 × g for 10 min, leaving the final pellets in ∼100 μL. DNA was extracted from the final pellet as described above.

Kidney: tissues were acquired from healthy wild-type Wistar rats. Fifty microliters of serial 10- fold dilutions of leptospires were spiked into 25 mg of kidney before DNA extraction to achieve concentrations of 1 × 10^7^ to 1 × 10^0^ leptospires per gram of tissue (21). After spiking, the tissues were processed in the same day for DNA extraction, or frozen (A) at −20 °C, and (B) at −80 °C before DNA extraction.

Whole blood: leptospires were added to 1 mL of EDTA-anticoagulated whole blood from healthy wild-type Wistar rats to achieve a final concentration of 1 × 10^7^ leptospires/mL. After spiking, serial 10-fold dilutions of 1 × 10^6^ to 1 × 10^0^ leptospires/mL were processed for DNA extraction on the same day or frozen (A) at −20 °C, and (B) at −80 °C before DNA extraction as described before.

All samples were tested in duplicate and three independent runs using *lipL32* and 23S Real-Time PCR assays as described before.

### Analytical Specificity

A total of 61 DNA of different culture strains representing 61 leptospiral species (Table S1) were tested by *lipL32* and *23S* Real-Time PCR. We also performed both assays using DNA from *E. coli*, *Salmonella enterica*, *Staphylococcus aureus*, *Bacillus cereus*, and *Pseudomonas aeruginosa*. Furthermore, *in silico* analysis was performed across different genera to verify the specificity of primers and probes of *lipL32* and *23S* assays.

### Performance and Validation on Clinical Samples

The performance of the new *lipL32* and *23S* qPCR was done using the DNA of a total of 451 clinical samples and compared with the results obtained from the former *lipL32* Real-Time PCR assay routinely used for the diagnosis of *Leptospira* spp. (9) to calculate standard diagnostic sensitivity (DSe) and specificity (DSp) (31) .

We tested 86 samples of human serum, classified as 26 positive and 60 negatives (Table S3). From animals, we tested 356 clinical samples (Table S4): 57 from horse (uterine samples), 60 from canine (urine samples), 48 from wildlife (kidney samples), 186 from bovine (36 semen, 34 urine, 8 epididymides, 6 testicles, and 102 uterine samples), and 5 from ovine (urine samples). Animal samples were classified as 178 negative and 178 positives.

The discrepancies of results obtained between the different assays were evaluated by direct sequencing of specific PCR products. We partially amplified the *16S* gene by PCR with primers 16S LA (GGCGGCGCGTCTTAAACATG) and 16S LB (TTCCCCCCATTGAGCAAGATT) (32) followed by nested PCR with primers 16S LC (CAAGTCAAGCGGAGTAGCAA) and s16S RS4 (TCTTAACTGCTGCCTCCCGT) (33) as described previously (34). PCR products were cleaned and submitted for Sanger sequencing. Sequence data were analyzed with Geneious Prime analysis software (Dotmatics). Consensus sequences were compared with available sequences in the GenBank database using BLAST.

### Bayesian Latent Class Analysis

To circumvent the fact that the former *lipL32* (9) is not a perfect gold standard test, as described before, we evaluated the diagnostic performance of our new optimized/developed assays using a Bayesian Latent Class Analysis (35). We evaluated the performance of our new assays with two different approaches using a web-based application (http://mice.tropmedres.ac), as previously described (36): a three-test one-population and a two-test two-population method. We used results obtained with the optimized *lip32*- and new *23S*-qPCR assays performed here compared with the results obtained with the original *lipL32*-qPCR assay routinely used (9). For the three-test one population method we used all animal clinical samples and considered the P1 and P2 results. For the two-test two population method we used all clinical samples (humans and animals) and the P1 results for the optimized *lipL32*-qPCR assay and *23S*-qPCR.

## RESULTS

### Real-Time PCR design and *in silico* evaluation

Each target gene was properly analyzed for probe and primer design. The *lipL32* gene was selected as an appropriate gene target for the detection of P1 species because of its successful use on a previously designed and validated assay (9, 19, 27). To avoid mismatches with more recently discovered P1 species, our final forward primer (lipL32_F) was shifted by four nucleotides at the beginning, starting at position 43 (previously position 47) and two nucleotides at the end, finishing at position 62 (previously position 64), based on the previous optimized design (LipL32-47Fd) (27). Furthermore, we kept the change from a C nucleotide to a degenerate nucleotide M on position 54, which can bind to both A and C. This alteration was performed due to the presence of an A nucleotide in the 54 position of *L*. *kmetyi* and *L. stimsonii* (27). The probe (lipL32_P1) and reverse primer (lipL32_R) were kept the same as in the previously original publication (9) and the recent optimization (27), respectively. Following these optimization strategies, both the primer and the probe sequences were absent from any major mismatches among P1 species. The exception was for *L. tipperaryensis* with two mismatches in the forward primer, two mismatches in the probe, and one mismatch in the reverse primer, and *L. gorisiae* with three mismatches in the forward primer, five in the probe, and one in the reverse primer (Figure 1). *L. gorisiae* presented the lower percentage of similarity of the *lipL32* gene compared with other P1 pathogenic species. Because P2, S1, and S2 species lacked the regions targeted by both the forward primer and the P1 probe utilized by the *lipL32* assay, the assay design suggested high specificity for P1 species only.

**Figure 1:**
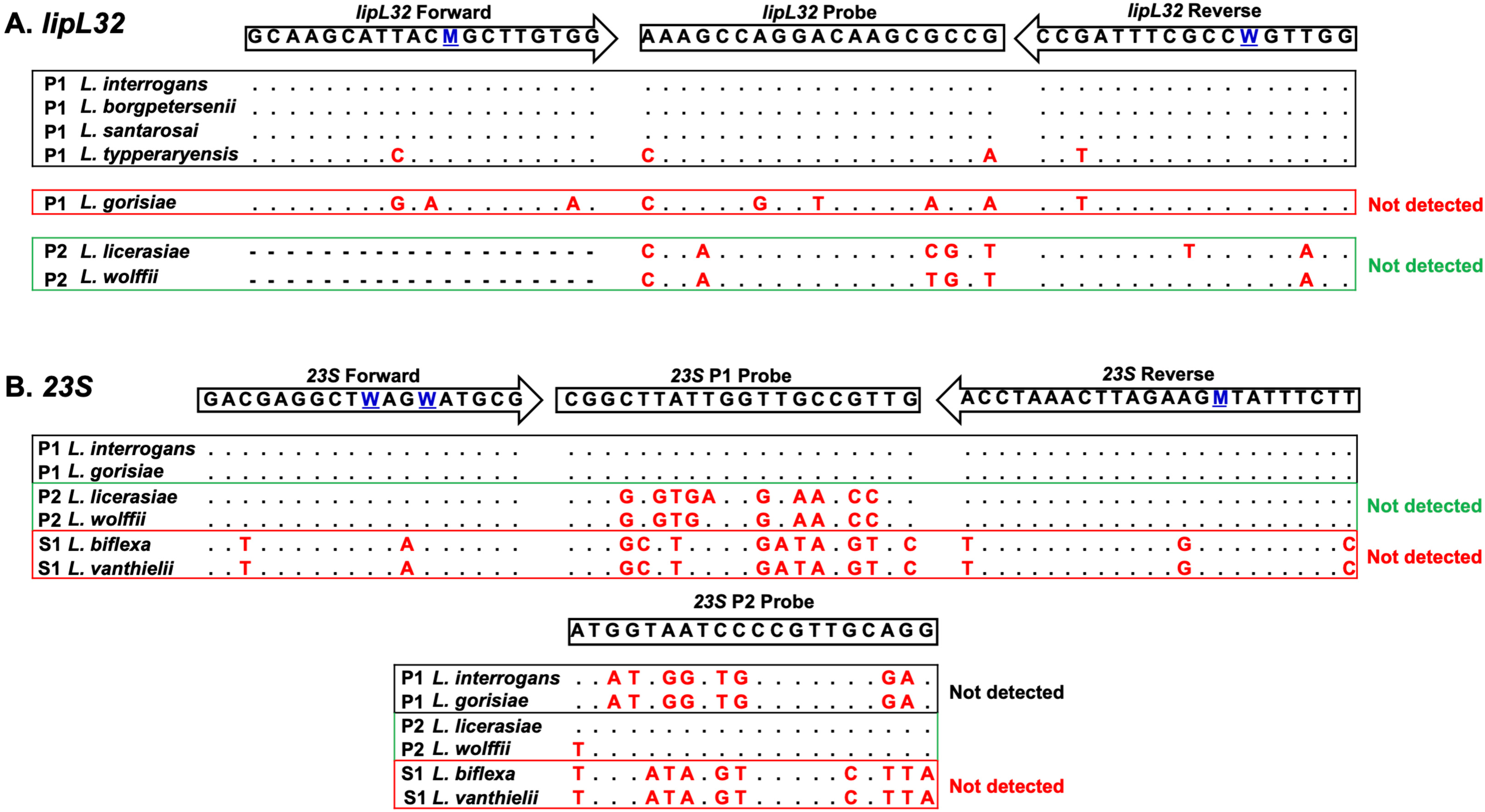
Sequence alignment for *lipL32* (A) and *23S* (B) primer/probe(s) set using representative species. The sequences of relevant representative species from P1, P2, and S1 clades were aligned to assess mismatches between primers and probes within species’ sequences. Aligned nucleotides are represented using a dot while dashes represent complete lack of alignment. Mismatches are represented by nucleotide codes in red. Degenerate nucleotide sequences are displayed in blue.

To identify an appropriate region for optimal group-specific probe design in the assay targeting the *23S rRNA* gene, we sought a target region of the *23S* sequence with high discrimination between P1, P2, and saprophytic species. The region of the P1-specific probe (23S_P1), on 1509-1528 position (LIC 10928), had an average of 10.94 and 12.96 mismatches with P2 and saprophytic species, respectively, while maintaining 100% homology among all P1 species. The P2-specific probe (23S_P2), on 1514-1533 position (AHOO02_rRNA1), had an average of 10.17 and 9.07 mismatches with P1 species and saprophytic species, respectively, while maintaining considerable homology among P2 species (Figure 1). Among P2 species, *L. perolatii, L. fluminis,* and *L. fletcheri* had 2 mismatched nucleotides with the P2 probe sequence, while *L. wolffii, L. broomii, L. inadai,* and *L. fainei* each had one mismatched nucleotide with the P2 probe sequence. Regions flanking the target probe region were screened for primer design with high homology between P1 and P2 species and low shared identity with all saprophytic species. The forward primer design (23S_F), on 1415-1432 (LIC 10928) and 1412-1429 (AHOO02_rRNA1) position, targeted a sequence of ∼100% identity between P1 and P2 species, as well as an average of 2.85 mismatches with saprophytic species, after an insertion of two degenerate nucleotide W (Figure 1). Similarly, the reverse primer design (23S_R), on 1630-1653 (LIC 10928) and 1631-1654 (AHOO02_rRNA1) position, targeted another region of ∼100% identity between P1 and P2 species and an average of 2.81 mismatches with saprophytic species, after an insertion of a degenerate nucleotide M (Figure 1).

### Calculation of Analytical Sensitivity and PCR Efficiency using LLOD

The serial dilutions of genomic DNA from *L. interrogans* serovar Copenhageni strain L1-130 were run on the *lipL32* assay with and without IPC (Figure 2A). The linear dynamic range (LDR) was 1 × 10^0^ to 1 × 10^7^ GEq/μL for both runs, with no statistical difference among them. The LLOD was 3.1 gene copies/reaction.

**Figure 2:**
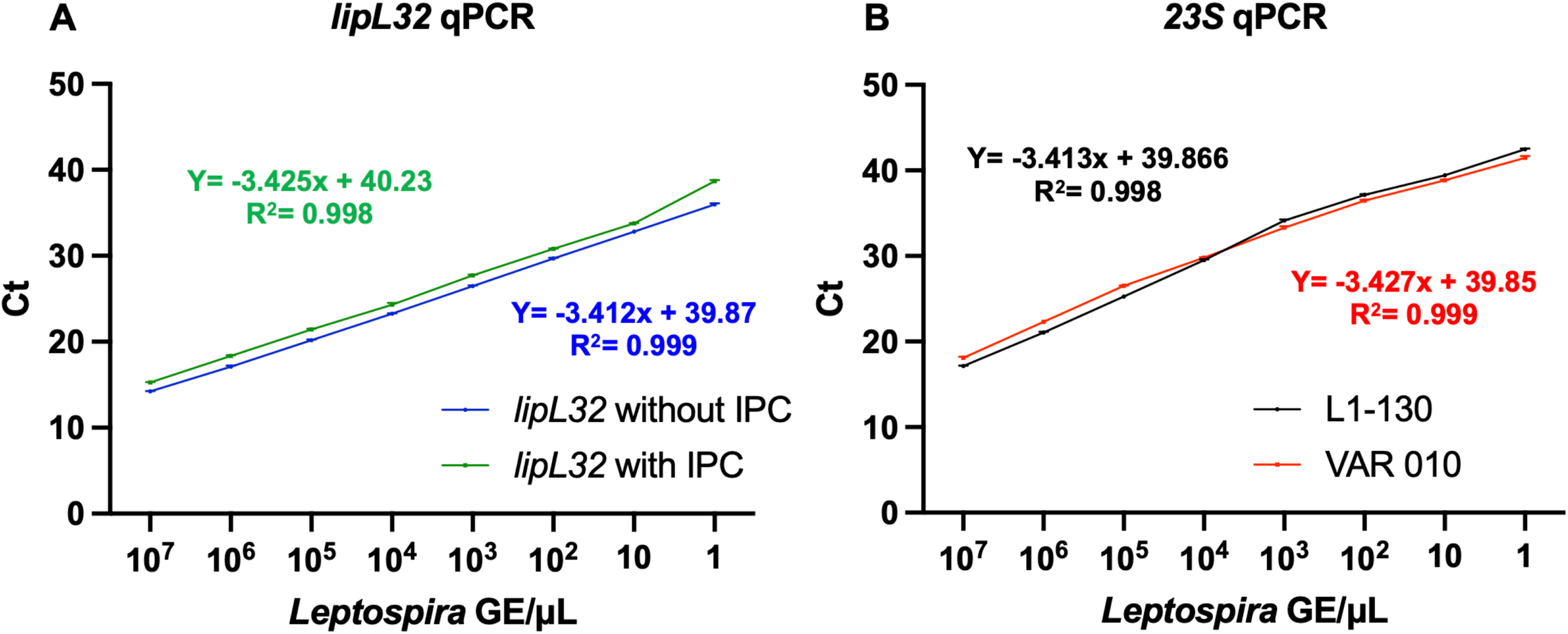
Lower limit of detection (LLOD) results for the *lipL32* real-time PCR assay using *L. interrogans* serovar Copenhageni strain L1-130 without IPC and with IPC (A) and for the *23S* real-time PCR assay using *L. interrogans* serovar Copenhageni strain L1-130 and *L. licerasiae* serovar Varillal strain VAR 010 (B). Error bars are measured as the SE but not visible.

The serial dilutions of genomic DNA from *L. interrogans* serovar Copenhageni strain L1-130 and *L. licerasiae* serovar Varillal strain VAR 010 (Figure 2B) were run on the *23S* assay. The linear dynamic range (LDR) was 1 × 10^0^ to 1 × 10^7^ GEq/μL for both species. The LLOD was 4.3 gene copies/reaction and 4.2 gene copies/reaction for pathogenic P1 and P2 respectively.

### Spiking Experiments

Urine: LLOD in urine, for *lipL32* and 23S assays, was 10 *Leptospira* GEq/mL when the sample was processed in the same day (A), same day after centrifugation at 900 × g for 10 min to remove potential debris (B), after 24h storage in ice (C), or after storage for 24 h in a Styrofoam container with ice packs followed by centrifugation to remove potential debris (D). After 48h storage in ice (E) or after storage for 48 h in a Styrofoam container with ice packs, followed by centrifugation to remove potential debris (F), the LLOD decreased to 10^2^ and 10^3^ *Leptospira* GEq/mL on the *lipL32* and *23S* assays respectively. Storage of the samples up to 48h without ice (G) or freezing the samples at −20 °C for 24 h without (H) or after centrifugation to remove potential debris (I) affected the LLOD of both assays negatively, reducing it to 10^3^ and 10^4^ *Leptospira* GEq/mL in *lipL32* and 23S assays, respectively (Table S2).

Kidney and whole blood: LLOD for tissues was 10 *Leptospira* GEq/mL in both assays irrespectively if the samples were processed on the same day or frozen for DNA extraction (Table S2).

### Analytical specificity and overall sensitivity of primers and probes

The analytical specificity for both assays was determined as 100%. Neither the *lipL32* nor *23S* assays detect any of the saprophytic *Leptospira* strains tested (Table 2). In addition, none of the other pathogenic organisms tested were detected by either assay. However, the optimized *lipL32* assay had a sensitivity of 95.2% with a CT average of 19.5. Despite the overall better efficiency and improved sensitivity, compared to the previous version of the assay, with 85.7% sensitivity and 24.4 CT average (Table 2), the mismatches identified in our *in-silico* analysis within the *L. gorisiae* species impacted the efficiency of the *lipL32* assay, preventing a positive detection. However, the mismatches identified within *L. tipperaryensis* didn’t affect the ability of the assay to identify this species, although the CT was higher (36.84) compared to the CT for all other species (Table 2). Nevertheless, the novel *23S* assay was not only 100% specific but also 100% sensitive for P1 and P2 pathogenic *Leptospira* when analyzing the results for the specific probes (Table 2).

**Table 2:** Results of *lipL32* and *23S* Real-Time PCR analysis of DNA representing 61 *Leptospira* species.

| Group | Species | Strain | Target and CTs |  |  |  |
| --- | --- | --- | --- | --- | --- | --- |
|  |  |  | *lipL32* 2009* | *lipL32* 2026<sup>#</sup> | 23S<sup>#</sup><br>P1 | P2 |
| P1<br>(21/21) | *L. adleri* | FH2-B-C1 | 21.4 | 15.14 | 15.51 | Undetermined |
|  | *L. ainazelensis* | 201903071 | 33.76 | 14.52 | 14.27 | Undetermined |
|  | *L. ainlahdjerensis* | 201903070 | 37.95 | 13.8 | 14.65 | Undetermined |
|  | *L. alexanderi* | L 60T | 33.35 | 20.83 | 25.8 | Undetermined |
|  | *L. alstonii* | 80-412T | 23.92 | 20.4 | 27.23 | Undetermined |
|  | *L. barantonii* | FH4-C-A1 | 19.5 | 15.09 | 14.98 | Undetermined |
|  | *L. borgpetersenii* | JB197 | 19.58 | 19.32 | 20.54 | Undetermined |
|  | *L. ellisii* | ATI7-C-A3 | 22.1 | 18.4 | 14.75 | Undetermined |
|  | *L. gomenensis* | KG8-B22 | 22.1 | 21.75 | 22.84 | Undetermined |
|  | *L. gorisiae* | WS92.C1 | Undetermined | Undetermined | 24.25 | Undetermined |
|  | *L. interrogans* | L1-130 | 18.87 | 15.96 | 24.96 | Undetermined |
|  | *L. kirschneri* | 3522C | 18.97 | 18.6 | 19.38 | Undetermined |
|  | *L. kmetyi* | Bejo-Iso9 | 23.58 | 20.73 | 20.6 | Undetermined |
|  | *L. mayottensis* | 200901122 | 29.5 | 20.24 | 23.98 | Undetermined |
|  | *L. noguchii* | CZ 214 K | 19.08 | 20.35 | 21.2 | Undetermined |
|  | *L. sanjuanensis* | LGVF01 | 32.5 | 24.76 | 28.17 | Undetermined |
|  | *L. santarosai* | 1342 K | 19.48 | 19.2 | 20.55 | Undetermined |
|  | *L. stimsonii* | Yale | Undetermined | 13.79 | 18.4 | Undetermined |
|  | *L. tipperaryensis* | GWTS #1 | Undetermined | 36.84 | 28.33 | Undetermined |
|  | *L. weilii* | Cox | 20.36 | 20.4 | 27.25 | Undetermined |
|  | *L. yasudae* | F1 | 23.83 | 20.52 | 24.2 | Undetermined |
| P2<br>(18/22) | *L. andrefontaineae* | PZF11-2 | Undetermined | Undetermined | Undetermined | 28.69 |
|  | *L. broomii* | 5399 | Undetermined | Undetermined | Undetermined | 30.85 |
|  | *L. cinconiae* | WS58.C1 | Undetermined | Undetermined | Undetermined | 21.81 |
|  | *L. dzoumogneensis* | M11A | Undetermined | Undetermined | Undetermined | 28.66 |
|  | *L. fainei* | But 6 | Undetermined | Undetermined | Undetermined | 31.24 |
|  | *L. fluminis* | SCS5 | Undetermined | Undetermined | Undetermined | 28.18 |
|  | *L. haakeii* | ATI7-C-A4 | Undetermined | Undetermined | Undetermined | 29.57 |
|  | *L. hartskeerlii* | MCA2-B-A3 | Undetermined | Undetermined | Undetermined | 29.33 |
|  | *L. inadai* | 10T | Undetermined | Undetermined | Undetermined | 32.54 |
|  | *L. koniamboensis* | TK1-4 | Undetermined | Undetermined | Undetermined | 26.27 |
|  | *L. licerasiae* | VAR 010 | Undetermined | Undetermined | Undetermined | 25.19 |
|  | *L. neocaledonica* | ES4-C-A1 | Undetermined | Undetermined | Undetermined | 29.64 |
|  | *L. perolatii* | FH1-B-B1 | Undetermined | Undetermined | Undetermined | 31.53 |
|  | *L. saintgironsiae* | FH4-C-A2 | Undetermined | Undetermined | Undetermined | 32.64 |
|  | *L. sarikeiensis* | LIMR175 | Undetermined | Undetermined | Undetermined | 29.54 |
|  | *L. semungkisensis* | SSS9 | Undetermined | Undetermined | Undetermined | 34.84 |
|  | *L. venezuelensis* | CLM-U50 | Undetermined | Undetermined | Undetermined | 23.21 |
|  | *L. wolffii* | Khorat-H2 | Undetermined | Undetermined | Undetermined | 33.63 |
| S1<br>(22/26) | *L. bandrabouensis* | M10A | Undetermined | Undetermined | Undetermined | Undetermined |
|  | *L. biflexa* | Patoc I | Undetermined | Undetermined | Undetermined | Undetermined |
|  | *L. bourretii* | PZF7-6 | Undetermined | Undetermined | Undetermined | Undetermined |
|  | *L. bouyouniensis* | M1A | Undetermined | Undetermined | Undetermined | Undetermined |
|  | *L. brenneri* | JW2-C-A2 | Undetermined | Undetermined | Undetermined | Undetermined |
|  | *L. congkakensis* | SCS9 | Undetermined | Undetermined | Undetermined | Undetermined |
|  | *L. ellinghausenii* | E18 | Undetermined | Undetermined | Undetermined | Undetermined |
|  | *L. harrisiae* | FH2-B-A1 | Undetermined | Undetermined | Undetermined | Undetermined |
|  | *L. iowaensis* | WS39.C2 | Undetermined | Undetermined | Undetermined | Undetermined |
|  | *L. jelokensis* | L5S1 | Undetermined | Undetermined | Undetermined | Undetermined |
|  | *L. kanakyensis* | TK5-11 | Undetermined | Undetermined | Undetermined | Undetermined |
|  | *L. levettii* | MCA2-B-A1 | Undetermined | Undetermined | Undetermined | Undetermined |
|  | *L. meyeri* | Went 5 | Undetermined | Undetermined | Undetermined | Undetermined |
|  | *L. mgodei* | WS4.C2 | Undetermined | Undetermined | Undetermined | Undetermined |
|  | *L. milleri* | WS60 | Undetermined | Undetermined | Undetermined | Undetermined |
|  | *L. montravelensis* | PZF5-3 | Undetermined | Undetermined | Undetermined | Undetermined |
|  | *L. mtsangambouensis* | M2A | Undetermined | Undetermined | Undetermined | Undetermined |
|  | *L. noumeaensis* | PZF14-4 | Undetermined | Undetermined | Undetermined | Undetermined |
|  | *L. perdikensis* | HP2 | Undetermined | Undetermined | Undetermined | Undetermined |
|  | *L. terpstrae* | ATCC 700639 | Undetermined | Undetermined | Undetermined | Undetermined |
|  | *L. vanthielii* | ATCC 700522 | Undetermined | Undetermined | Undetermined | Undetermined |
|  | *L. wolbachii* | CDC | Undetermined | Undetermined | Undetermined | Undetermined |

\* Stoddard et al, 2009.

### Clinical validation

Using the DNA from human clinical samples (serum), sensitivity and specificity for *lipL32* and *23S* were calculated as 100% (Tables S3). With the DNA from animal samples, *lipL32* and *23S* assays had a sensitivity of 100% and specificity of 89.9% and 89.3%, respectively (Tables 3 and S4). However, it is important to emphasize that both the optimized *lipL32* and the new 23S assays determined as positives the same 18 samples that were negative in the previous *lipL32* assay. Furthermore, one negative sample, dog urine, although negative on all assays regarding P1 species, was determined positive when evaluating the novel P2 probe of the *23S* assay (Table S4).

To better understand and analyze this inconsistency, we performed a Bayesian latent class model (Bayesian LCM), which assumes that there is no perfect assay. For reliability of the results, as required by the models, all parameters were obtained with a high level of convergence of chain and high fitness of frequencies predicted by the Bayesian LCM with the observed data. In our three-test one-population method, we focused on all the animal clinical samples that generated the inconsistencies mentioned above. The Bayesian LCM for the optimized *lipL32* assay described here showed a 99.9 % for diagnostic sensitivity (DSe) and specificity (DSp), and also for positive (PPV) and negative predictive value (NPV), in contrast with a DSe of 90.7% and a NPV of 89.8% for the original *lipL32* (9) (Table 3). The DSe and NPV for the *23S* assay was also high (99.9%), however the DSp and PPV were affected (99.3% and 99.4%, respectively) since we considered the P2 positive result on this assay (Table 3). Nevertheless, when performing the Bayesian latent test using a two-test two-population method (P1 results for human and animal clinical samples), the new *lipL32* and *23S* assays showed a much better performance compared to the original *lipL3*2 assay (Table S5).

We also performed molecular analysis of the samples that had inconsistency results. All 19 samples were amplified by 16S, and PCR products gave interpretable sequences. Of the 18 analyzed sequences (positives by the new *lipL32* and *23S* P1), six (33.3%) were identified as *L. alstoni* with 99.5% of similarity, and twelve (66.7%) as *L. santarosai* with 98.8% of similarity. The analyzed sequence that was positive by the *23S* P2 assay, the results indicated *L. wolffii* with 98.65% of similarity. Taken together, those results indicated an improved efficiency of the novel assays to identify positive samples, including the potential role of P2 species on infection.

**Table 3.**
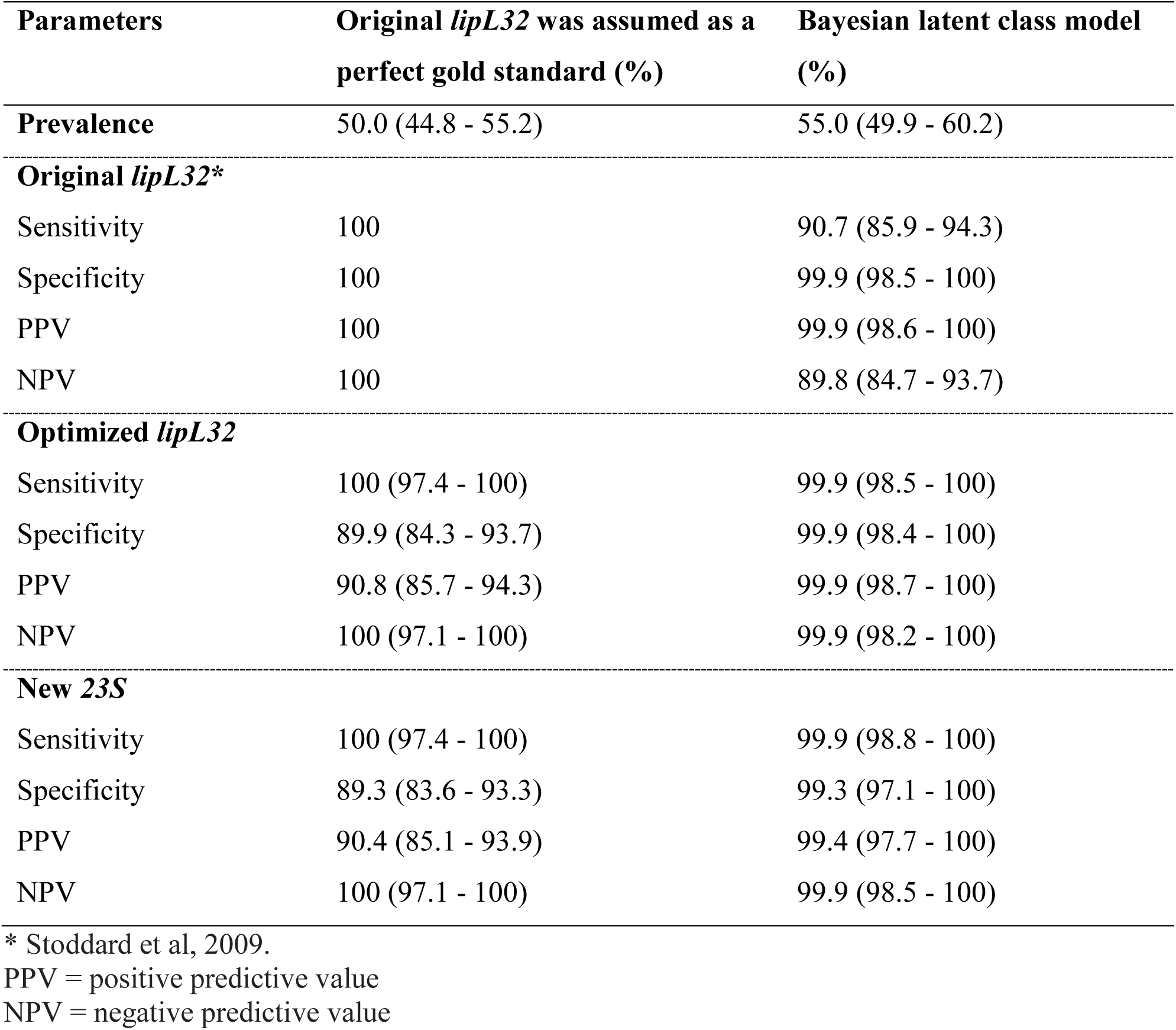
Diagnostic performance of lipL32 and 23S-based qPCR comparing 1 traditional evaluation methods and Bayesian latent class model using a three-test one population method. Values shown are estimated means with 95% confidence interval.

## DISCUSSION

Leptospirosis is the most widespread reemerging zoonosis globally and has the potential to threaten livestock supply and medical infrastructure of lower- and middle-income countries. However, there is a limited body of literature that describes the clade- and species-specific virulence and environmental survival of *Leptospira*. Thus, recent developments in molecular assays that can detect and differentiate between *Leptospira* species presenting in various clinical and environmental contexts have provided increased opportunities to investigate infection and transmission properties of *Leptospira* species (5, 9). When considering the need for improved assays that may be used in various epidemiological contexts, Real-time PCR (qPCR) assays offer the potential for rapid, sensitive, and group-specific diagnostics that can discern clades of *Leptospira* present and thriving in certain epidemiological environments and/or animal hosts. The increased use of qPCR-based assays has the potential to improve the underreporting of leptospirosis considering challenges with diagnostics such as cost, time, and sensitivity, especially in lower-income settings (37). In this study, we developed, optimized, and validated two qPCR-based assays targeting different genes with specific but additive results, which can be used individually or in combination to improve leptospiral identification.

Most human and animal cases of leptospirosis are caused by species of the P1 clade, and several animal models have also identified this clade as the most pathogenic species (4). A real-Time qPCR specific for *lipL32*, a conserved gene unique to pathogenic leptospires that discriminates from saprophytic species (17, 18), remains the most promising target for PCR-based detection of species of the P1 clade (13). The recent discovery of several new P1 species (5–7), many identified after the development of the first clinically validated TaqMan qPCR assay targeting the *lipL32* gene (9, 13), highlights the need to improve the assay’s efficiency (27). Previous attempts to refine the assay, although successful at the time, have been similarly influenced by ongoing identification of new species (9, 13, 19, 27). The present study builds on these prior approaches by incorporating available *lipL32* sequences from newly described P1 species, creating an opportunity to further enhance the assay’s specificity and sensitivity.

The *23S*-based assay developed in this study utilizes a multiplex design to simultaneously detect and differentiate between P1 and P2 *Leptospira* species using a single set of primers. 23S ribosomal RNA molecules have been discussed as promising molecular candidates to differentiate between P1 and P2 species, based on their considerable conservation among P1 and P2 species but sequence divergence in saprophytic species (10). Within the phylogenetic division of the *Leptospira* genus into four clades, the species belonging to both P1 and P2 clades are considered infectious (5). These species can cause disease in various mammal species, including humans (38), although for some, clinical or laboratory evidence remains limited. Different species from the P2 clade have been found in febrile cases in humans (39–44) and as the cause of disease in different animals (43, 45, 46). These findings highlight the relevance of studying the P2 clade, since its role in the dynamics of epidemiology and maintenance of the disease, as well as the link between different host species and humans, remains unclear (5).

Pathogenic leptospires are only detectable in the blood during the first week after onset of symptoms, and animal reservoirs might maintain leptospires in their kidneys without showing any symptoms (2, 4). With a LLOD of 10 *Leptospira* GEq/mL in blood samples, the *lipL32*- and *23S*-based assays could be useful tools for the rapid diagnosis of acute leptospirosis in humans and animals, particularly in severe cases such as severe pulmonary hemorrhagic syndrome (SPHS) in humans (2), where serology or culture are not fast enough to guide clinically decision-making. Although the shedding of leptospires in urine can be intermittent, urine is a useful sample for the confirmation of leptospirosis, especially in animals. Our results showed that the *lipL32* and *23S* assays were able to detect low concentrations of leptospires (1 *Leptospira* GEq/mL) when testing urine samples processed on the same day or after storage for 24 h in ice, with or without centrifugation. Storage of the samples for 48 h with or without ice packs or after freezing, with or without centrifugation, affected the LLOD negatively in both assays. However, the negative effect remained one log higher in the *23S*-based assay, indicating a better sensitivity of the *lipL32* assay depending on the storage time and method of urine samples. There was no effect on the LLOD when using blood or kidney tissue, with or without freezing before processing of the samples. Additionally, we demonstrated a high specificity of the *lipL32* and *23S* Real-Time qPCR for pathogenic *Leptospira*, with no cross-reaction with other common pathogens or unspecific reactions from blood, tissue, or urine samples.

Bayesian methods assume that all tests evaluated are imperfect and have been often used to assess the reliability of diagnostic tests in the absence of a true gold standard, including estimation of prevalence and diagnostic sensitivity and specificity using samples with unknow status (35, 37). When assuming the original *lipL32* assay as a “perfect reference test”, our clinical evaluation of both assays showed 100% sensitivity and specificity with serum samples of human with suspected leptospirosis. However, the evaluation with clinical samples from animals showed a high sensitivity but lower specificity for the optimized *lipL32* and novel *23S* assays. Both assays were able to detect more positive samples, compared to the prior *lipL32* assay, including a sample positive for a P2 species, when using the *23S*-specific probe. Using a Bayesian LCM analysis, together with sequencing confirmation of the samples with discrepancies, we demonstrated a higher sensitivity and specificity for the new assays described here, emphasizing the advantages of assays that are designed targeting all species of pathogenic *Leptospira*.

Both assays described here demonstrated a high analytical specificity and sensitivity with an improved DSe and DSp when compared to the former *lipl32*-qPCR assay (5, 9). However, when evaluating different *Leptospira* species detection, our *lipL32*-based assay failed to identify all P1 pathogenic species as expected. Recently isolated from a water source in Iowa, United States (7), the *L. gorisiae* P1 species showed a surprisingly low identity for its *lipL32* gene, 83%, in contrast to an average of 90% identify among other P1 species when comparing to the *L. interrogans* strain Fiocruz L1-130 *lipL32* sequence. This major difference jeopardized the ability to optimize the *lipL32* primers and probe to enable the detection of this specific species. Nevertheless, our *23S*-based assay was able to detect all P1 species, including *L. gorisiae*, and all P2 species with their specific probes. Despite the efficiency and simplicity of the *lipL32*-based assay, it is essential to highlight that the *23S* assay is an important alternative for detecting all pathogenic species of *Leptospira* in clinical samples. Thus, combining these assays may present opportunities for the development of a new method of clinical diagnosis of leptospirosis among mammal hosts and reservoirs. Although these assays were not evaluated on environmental samples in this study, we believe that either assay could also be useful in better understanding the etiology and epidemiology of leptospirosis in the environment, and we are currently conducting studies to validate this hypothesis. Whether the assays are most effective alone or in combination, will be based on expected outcomes, the sample types, and how quickly the samples can be processed.

Focusing on the One Health approach, increasing the capacity for the detection of *Leptospira* in humans, animals, and environmental samples will improve our efforts on public health surveillance while providing alternatives to control and prevent cases of leptospirosis in animals and humans. These newly optimized and developed assays resulted in a lower limit of detection and increased diagnostic sensitivity and specificity, while retaining the ability to detect and differentiate between P1 and P2 pathogenic species of *Leptospira*. Both assays will improve detection of clinical infections during early stages, investigation of urinary shedding and renal carriage rates, and support future studies on the ecoepidemiology of animal and human leptospirosis. Thus, these assays may ultimately help to guide treatment and interventions to prevent zoonotic spillover off this important neglected tropical disease.

## DATA AVAILABILITY

All data are available in the manuscript.

## ETHICS APPROVAL

Animal protocol was approved by the Institutional Committee for the Use of Experimental Animals, University of Connecticut (protocol no. A26-023). The use of human samples was approved by the Oswaldo Cruz Institute Ethics Committee (protocol no. 74156723.2.0000.5248)

## Data Availability

All data produced in the present work are contained in the manuscript.

## ACKNOWLEDGEMENTS

This research was supported in part by NIH grants R01AI182354 and R21AI163663 (EAWJ), and USDA capacity grant H7007836 (EAWJ). We thank Dr. Jarlath Nally (ARS, USDA) for providing strains JB197 and GWTS#1, and Dr. Mathieu Picardeau (Institute Pasteur, France) for providing DNA from different *Leptospira* species. We thank Liana Barbosa (Wunder Lab) for her technical work. We thank CNPq (444137/2024-6) for which WL is a fellow.

---

**Table S1:** Accession number, DNA availability, and references for all 74 species of Leptospira used in this study.

**Table S2:** Lower limit of detection (LLOD) of *lipL32* and *23S* qPCR in spiked clinical samples. Samples were spiked with different concentrations of pathogenic *Leptospira* (10^7^ – 10^0^) before DNA extraction, qPCR, and LLOD analysis.

**Table S3:** Validation of *lipL32* and *23S* Real-time PCR assay for pathogenic *Leptospira* detection in DNA of clinical human samples.

**Table S4:** Validation of *lipL32* and *23S* Real-time PCR assay for pathogenic *Leptospira* detection in DNA of clinical animal samples.

